# Chronic Kidney Disease Prediction Using Neural Approach

**DOI:** 10.1101/2020.06.28.20142034

**Authors:** Shawni Dutta, Samir Kumar Bandyopadhyay

## Abstract

All over the world, chronic kidney disease (CKD) is a serious public health condition that needs to be detected in advance so that costly end-stage treatments like dialysis, kidney transplantations can be avoided. Neural network model and 10-fold cross-validation methodology under a single platform in proposed as well as implemented in order to classify patients with CKD. This will assist medical care fields so that counter measures can be suggested. The performance of proposed classifier is justified against other baseline classifiers such as Support Vector Machine, K-Nearest Neighbours, Decision tree and Gradient Boost classifier. Experimental results conclude that the performance of neural network with 10-fold cross-validation method reaches promising accuracy of 98.25%, f1-score of 0.98, and kappa score of 0.96 and MSE of 0.0175.

## Introduction

CKD is often responsible for accelerating as an interfering cause of Cardiovascular Disease (CVD). CKD is associated with several parameters such as age, diabetes, hypertension, obesity, primary renal disorders [1]. In India, CKD appears to be frequently increasing and at least 70% people in rural areas suffer from CKD which is often diagnosed at later stages. During the last stage less than 10% of renal disease patients can afford any kind of renal replacement therapy due to quiet substantially high cost of treatment [2]. The primary function of the kidney of CKD patients suffers from blood filtration gradually over time. Regular hemodialysis or kidney transplant are required to survive at the end stage. During the development of disease, patients suffer from complications of acidosis, anemia, diabetes, high blood pressure or neuropathy. It affects patients’ quotidian life. The median survival time of late-stage patients is very short. It is importance to check up patient within short period of time since it helps to decide appropriate care, medication and medical interventions of the patient. It creates a complex interrelationship and influence the outcome of the individual patient. A prediction model is able to fit into that role and may be used to revise current treatment. Due to the complex nature of the problem and multiple interrelated factors may influence the patient’s survival, such a model is a challenging task.

Data mining and knowledge discovery processes allow health records to be used systematically so that hidden patterns among enormous database can be identified [3]. This will help health care industries for obtaining efficient treatment, eliminating risks from patients’ life and maintaining low amount of medical expense. Data mining techniques are useful in health care systems in terms of predicting diseases in advance as well as assist the doctors to take precaution measure in case of diagnosis process. Machine Learning (ML) methods fulfils the objective of detecting CKD at an early stage by analysis all related factors those have impact on CKD. Classification is a supervised machine learning technique that analyses specified set of features and identifies data as belonging to a particular class. This research aims to identify patients with CKD by analysing interfering factors such as diabetic tendency, blood reports and many more. In this paper, neural network [4] followed by 10-fold stratified cross-validation methodology [5] is implemented in this paper as an automated tool for CKD prediction. This paper also implements Support Vector Machine (SVM) [6], K-Nearest Neighbours (k-NN) [7], Decision Tree (DT) [8], Gradient Boost algorithm [9] and compares the performance of proposed method with these classifiers.

## Related Works

Numerous researches have been carried out that aim to predict CKD. Using four machine learning approaches such as, RPART, LogR, SVM, MLP have been applied to CKD dataset and 0.995 of AUC was obtained as highest score [10]. Another study explored kidney function failure using classification algorithms such as Back Propagation Neural Network, Radial Basis Function and Random Forest. These algorithms are evaluated against dataset from the Coimbatore state for about 1000 patients with 15 attributes. Experimental results concluded that the radial basic function to be the best classifier with 85.3% prediction accuracy [11].

For determining severity stage in chronic kidney disease, the results of applying Probabilistic Neural Networks (PNN), Multilayer Perceptron (MLP), SVM and Radial Basis Function (RBF) algorithms have been compared. Experimental results show that the PNN algorithm provides better classification and prediction performance [12]. Another study employed SVM and ANN in order to predict Kidney disease. After comparing the performance of these two predictive models, it has been concluded that ANN outperforms SVM with an accuracy of 87% [13].

## Dataset Used

This paper collects *Chronic Kidney Disease Data Set* from UCI machine learning repository [20]. The dataset contains 400 numbers of records and each record is formulated as collection of 26 variables. Information regarding these 25 attributes is provided in Table 1. The attribute ‘classification’ variable identifies whether the patient has CKD or not. This variable is kept as dependent or target variable during classification process. The rest variables are fed as input to classifier model in order to predict the target class. The distribution of the target variable in the dataset is depicted in fig1.

**Table 1.** Description of Collected dataset.

| Attribute Name | Description | Type of Attribute | Values Present | Count of Missing Values |
| --- | --- | --- | --- | --- |
| Age | Patients' age | numerical | 2-90 | 9 |
| Bp | Blood pressure in mm/Hg | numerical | 50-100 | 12 |
| Sg | Specific gravity | nominal | 1.005-1.025 | 47 |
| Al | Albumin | nominal | 0-5 | 46 |
| Su | Sugar | nominal | 0-5 | 49 |
| Rbc | Red blood cells | nominal | Normal, abnormal | 152 |
| Pc | Pus cell | nominal | Normal, abnormal | 65 |
| Pcc | Pus cell clumps | nominal | Present, not present | 4 |
| Ba | bacteria | nominal | Present, not present | 4 |
| Bgr | blood glucose random in mgs/dl | numerical | 22.0-490.0 | 44 |
| Bu | blood urea in mgs/dl | numerical | 1.5-391.0 | 19 |
| Sc | serum creatinine in mgs/dl | numerical | 0.4-76.0 | 17 |
| Sod | Sodium in mEq/L | numerical | 4.5-163.0 | 87 |
| Pot | Potassium in mEq/L | numerical | 2.5-47.0 | 88 |
| Hemo | Haemoglobin in gms | numerical | 3.1-17.8 | 52 |
| Pcv | packed cell volume | numerical | 9-54 | 70 |
| Wc | white blood cell count | numerical | 3800-21600 | 105 |
| Rc | red blood cell count | numerical | 2.3-8.0 | 130 |
| Htn | hypertension | nominal | Yes, no | 2 |
| Dm | diabetes mellitus | nominal | Yes, no | 2 |
| Cad | coronary artery disease | nominal | Yes, no | 2 |
| Appet | appetite | nominal | Good, poor | 1 |
| Pe | pedal edema | nominal | Yes, no | 1 |
| Ane | anaemia | nominal | Yes, no | 1 |
| Classification | Patients having CKD or not | nominal | Ckd, non-Ckd | 0 |

**Fig. 1:**
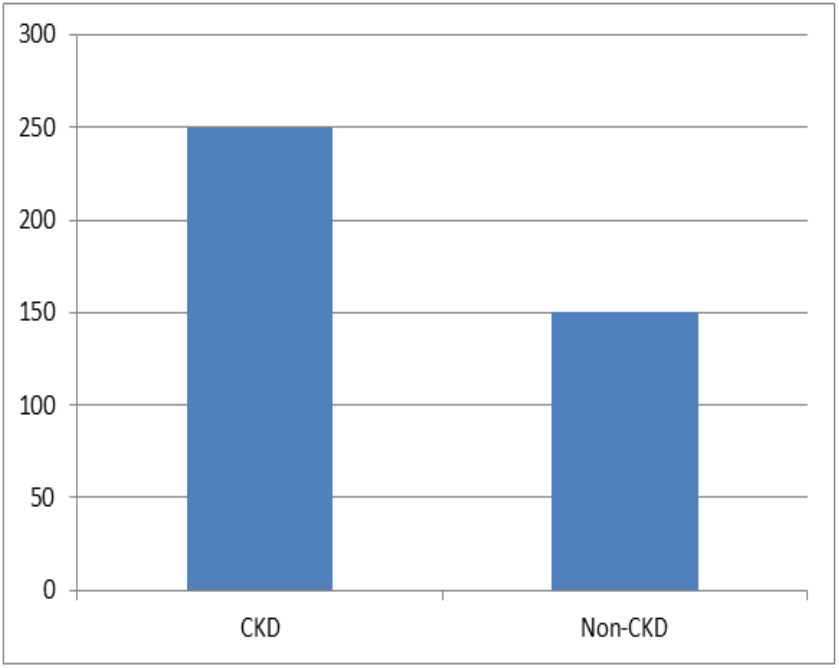
Distribution of target variable on the dataset.

Once the data collection is completed, it is necessary to put some concentration on missing values present in the dataset. Presence of missing values will change the prediction efficiency. However, the presence of missing values can be ignored or deleted when the number of missing values is less in percentage. In some cases, it is required to consider unknown or missing values present in the dataset since these may contribute to the disease. In our implementation, missing values are handled by replacing mean or average value of considered attribute. This will assist to obtain more accurate and genuine prediction results. After this, present nominal variables are converted to numerical values of range 0 to 1. These steps will assist to obtain pre-processed dataset. This dataset can now be fitted to any classifier model.

## Proposed Methodology

In this framework, neural network model is built and next 10-fold stratified cross-validation methodology is implemented as classifier model for identifying patients with CKD. The target of this classification is to identify whether a patient can be affected by CKD or not. Neural networks mimic the operation of human brains and recognises underlying relationship among the data. Neural network model is constructed by assembing multiple layers with linear or non-linear activation functions. These layers are trained together to learn for a complex problem solving approach [4]. Activation functions are capable of executing complex computations and produce outputs within a certain range. In other words, activation function is a step that maps input signal into output signal [14]. ‘relu’ and ‘sigmoid’ are two popular activation functions that are explained as follows

- Sigmoid activation function [14] transforms input data in the range of 0 to 1 and it is shown in equation(1).

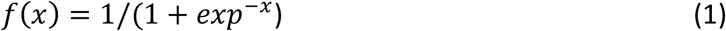
- Relu activation function [14] is a faster learning Activation function which is the most successful and widely used function. It performs a threshold operation to each input element where values less than zero are set to zero whereas the values greater or equal to zeros kept as intact and it is shown in equation (2).

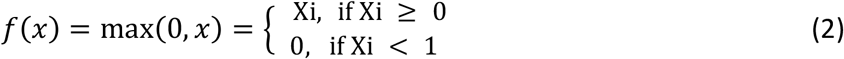

The neural network is implemented by incorporating three dense layers by adding number of nodes 128,32,1 respectively. These layers are activated by either ReLu or Sigmoid function. These layers are compiled using ‘adam’ [15] solver. Using ‘binary cross entropy’ loss function is endeavored as training criterion. This model is trained through 30 epochs and with the batch size of 10. Fine-tuning these hyper-parameters will enhance the performance of this model. During training phase, this model receives 7,361 parameters and trains those parameters. Detailed description of this model in terms of layers used, number of units attached, shape of output from each layer, activation function used in each layer is provided in Table2.

Once the model is constructed, 10-fold stratified cross-validation methodology is implemented. It is a resampling methodology where the dataset is segregated into 10 groups and in each iteration one group is considered as the test data and the remaining nine folds are considered as training data. The above mentioned model is fitted into the training dataset and it is evaluated against the test dataset. Later evaluation scores for each of these iterations are accumulated and mean score is calculated. The implementation of cross-validation ensures stratified mechanism which enforces that the distributions of all folds are necessarily similar to proportion of all labels in the original data [5].

### Baseline Classifiers

This section elaborates implementation of other classifiers such as K-NN, SVM, DT, Gradient Boost algorithms. Brief description of all these classifier models along with their implementation is explained in this section.

Support Vector Machine (SVM) [6] is quite advantageous in handling classification tasks with superior generalization performance. The method minimizes the upper limit of the generalization error based on the structural risk minimization principle. SVM can map input vector to a higher dimensional space by constructing a maximal separating hyper-plane. Two parallel hyper-planes are constructed on each side of the hyper-plane that separate the data. The separating hyper-plane is the hyper-plane that maximizes the distance between the two parallel hyper-planes. In order to obtain better generalization error by the classifier, the maximised distance between these parallel hyper-planes are considered [6].

K-Nearest Neighbours (K-NN) [7] is a supervised ML algorithm that is often known as Memory based classification. During classification process, it considers identifies objects based on closest proximity of training examples in the feature space. It is known as lazy learners because during training phase it just stores training samples. The classifier considers k number of objects as the nearest object while determining the class. The main challenge of this classification technique relies on choosing the appropriate value of k. While calculating distance among instances, Minkowski distance [7] is considered to be general metric for any data.

A Decision Tree (DT) [8] is a classifier that exemplifies the use of tree-like structure. Each target class is denoted as a leaf node of DT and non-leaf nodes of DT are used as a decision node that indicates certain test. The outcomes of those tests are identified by either of the branches of that decision node. Classification results are obtained by starting from the beginning at the root this tree are going through it until a leaf node is reached [8].

Gradient Boost algorithms [9] are suitable in fitting new models to provide maximised efficiency while estimating response variable. The objective of this algorithm is to construct new base learners to be maximally correlated with the negative gradient of the loss function, associated with the whole ensemble. This algorithm is highly customizable to any domain which provides freedom in model designing. One of the important issues of this algorithm is identifying and incorporating loss function to this algorithm which is subject to change as a matter of trial and error [9].

### Implementation of baseline classifiers

This section explains the implementation of SVM, k-NN, DT and gradient boost classifiers. The pre-processed data is divided with the ratio of 67:33 for obtaining training and testing dataset respectively. The training data is fitted into the classifier models and later prediction is collected for the testing dataset. The implementation of SVM classifier uses regularization parameter C= 1.0 and Radial basis function (RBF) as kernel function. The k-NN classifier receives value of k as 5. The Minkowski distance is used while calculating distances among instances. The Gradient Boost classifier is learnt with a rate of 1.0 and using 500 numbers of estimators for achieving the best known performance. The DT classifier is implemented using *Gini* index and *best* supportive algorithm as splitter. The prediction performances of these classifiers will help to justify the performance of the proposed classifier.

### Classifier Evaluation metrics

The performance of predictive models needs to be evaluated which instantiates the importance of evaluation metrics. This section discusses the metrics those are employed to measure the performance of the classifier models.

1. Accuracy [17] is a metric that detects the ratio of true predictions over the total number of instances considered. However, the accuracy may not be enough metric for evaluating model’s performance since it does not consider wrong predicted cases. Hence, for addressing the above specified problem, precision and recall is necessary to calculate.
2. Precision [18] identifies the ratio of correct positive results over the number of positive results predicted by the classifier. Recall denotes the number of correct positive results divided by the number of all relevant samples. F1-Score or F-measure [18] is a parameter that is concerned for both recall and precision and it is calculated as the harmonic mean of precision and recall. The best value of F1-score, precision, and recall is known to be 1.
3. Mean Squared Error (MSE) [18] is another evaluating metric that measures absolute differences between the prediction and actual observation of the test samples. MSE produces non-negative floating point value and a value close to 0.0 turns out to be the best one.
4. Cohen-Kappa Score [19] is also taken into consideration as an evaluating metric in this paper. This metric is a statistical measure that finds out inter-rate agreement for qualitative items for classification problem. The kappa statistic outputs value in the range of -1 to +1 and +1 indicates the maximum chance of agreement.

Precisely, the above mentioned metrics can be defined as follows with given True Positive, True Negative, False Positive, False Negative as TP,TN,FP,FN respectively-

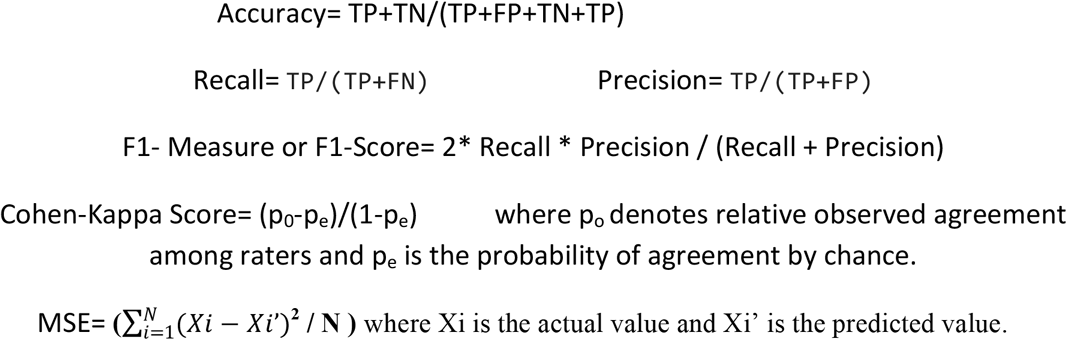

## Experimental Results

The training scores as well as testing scores obtained for each fold are depicted in Fig2. Accuracy, F1-Score, Cohen-kappa score and MSE were the scoring methods for measuring the performance of the proposed model. Once evaluation is done for all folds of cross-validation, mean score for testing dataset is calculated for each mentioned metrics. This implementation is validated against other baseline classifiers such as SVM, k-NN, DT and gradient boost algorithms in terms of aforementioned evaluation metrics. The comparative study is shown in Table 2. This analysis denotes that the proposed method is quite superior in terms of CKD detection with respect to baseline classifiers.

**Fig. 2.**
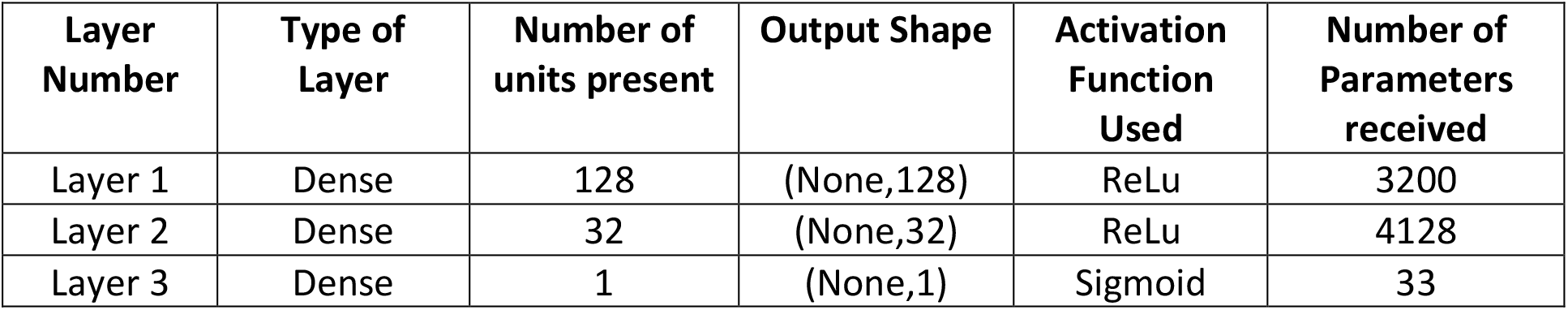
Summary of Neural Network Model.

**Fig. 2.**
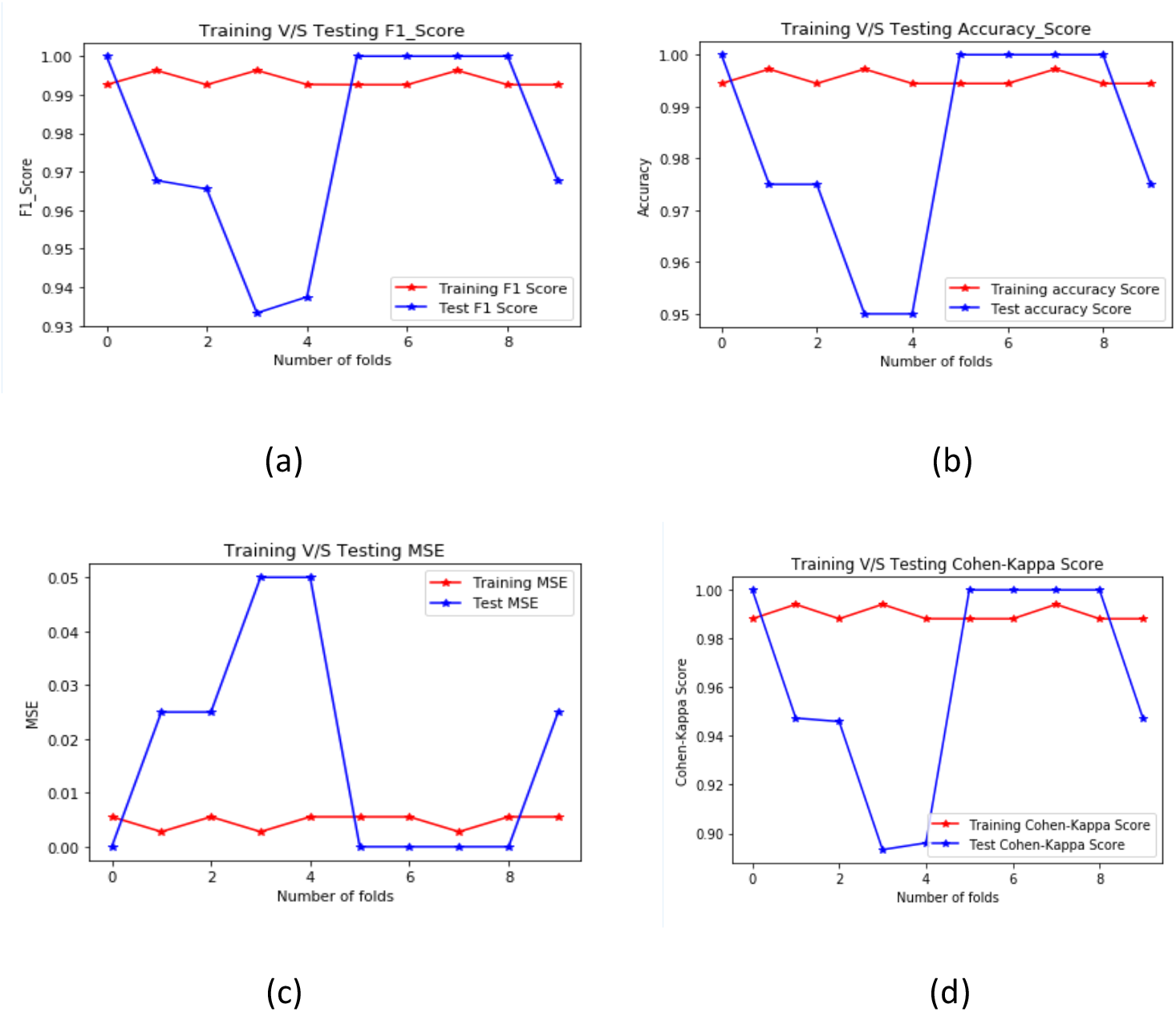
Training and Testing (a) F1-Score (b) accuracy score (c) MSE (d) Cohen-kappa score for each fold of cross-validation.

**Table 2.** Performance Comparison of Proposed model and Baseline classifiers.

| Classifiers-> | Proposed Classifier | Baseline Classifier |  |  |  |
| --- | --- | --- | --- | --- | --- |
| Performance Measure Metrics | 10-Fold Cross Validated Neural Network | SVM | K-NN | DT | Gradient Boost Classifier |
| Accuracy | 98.25% | 96.21% | 91.67% | 94.7% | 97.73% |
| F1-Score | 0.98 | 0.96 | 0.92 | 0.95 | 0.977 |
| Cohen-Kappa Score | 0.96 | 0.92 | 0.83 | 0.89 | 0.95 |
| MSE | 0.0175 | 0.0379 | 0.0833 | 0.053 | 0.0227 |

## Conclusions

CKD classification and detection may be used as analyzing tool for giving the second opinion to the doctors and pathologists. This paper proposed an approach for the prediction of chronic kidney disease using machine learning techniques. In the proposed research neural network model is initially built and next 10-fold stratified cross-validation methodology is implemented as classifier model for identifying patients with CKD. The classification identifies whether a patient can be affected by CKD or not.

## Data Availability

The data is available in the INternet.

http://archive.ics.uci.edu/ml

